# *serosim*: an R package for simulating serological survey data arising from vaccination, epidemiological and antibody kinetics processes

**DOI:** 10.1101/2023.01.24.23284958

**Authors:** Arthur Menezes, Saki Takahashi, Isobel Routledge, C. Jessica E. Metcalf, Andrea L. Graham, James A. Hay

## Abstract

*serosim* is an open source R package designed to aid inference of serological surveys, by simulating data arising from user-specified vaccine and infection-generated antibody kinetics processes using a random effects model. Serological surveys are used to assess population immunity by directly measuring individuals’ antibody titers. They uncover locations and/or populations which are susceptible and provide evidence of past infection or vaccination to help inform public health measures and surveillance. Both serological surveys and new analytical techniques used to interpret them are increasingly widespread. This expansion creates a need for tools to simulate serological surveys and the processes underlying the observed titer values, as this will enable researchers to identify best practices for serological survey design, and provide a standardized framework to evaluate the performance of different inference methods. *serosim* allows users to specify and adjust model inputs representing underlying processes responsible for generating the observed titer values like time-varying patterns of infection and vaccination, population demography, immunity and antibody kinetics, and serological survey sampling design in order to best represent the population and disease system(s) of interest. This package will be useful for planning sampling design of future serological surveys, understanding determinants of observed serological data, and validating the accuracy and power of new statistical methods.

**Author Summary:** Public health researchers use serological surveys to obtain serum samples from individuals and measure antibody levels against one or more pathogens. When paired with appropriate analytical methods, these surveys can be used to determine whether individuals have been previously infected with or vaccinated against those pathogens. However, there is currently a lack of tools to simulate realistic serological survey data from the processes determining these observed antibody levels. We developed *serosim*, an open source R package which enables users to simulate serological survey data matching their disease system(s) of interest. This package allows users to specify and modify model inputs responsible for generating an individual’s antibody level at various levels, from the within-host processes to the observation process. *serosim* will be useful for designing more informative serological surveys, better understanding the processes behind observed serological data, and assessing new serological survey analytical methods.

## Introduction

Serological surveys, also known as serosurveys, measure individual biomarker quantities, namely antibody titers, across populations to help uncover important hidden epidemiological variables such as susceptibility and past epidemic and vaccination trends[1]. These hidden variables are required to predict and prevent outbreaks at the population level, and while they may be indirectly inferred via vaccination coverage and incidence data, this inference is subject to inaccuracies since vaccination coverage does not directly translate to individuals immunized and incidence data is often underreported and incomplete[2–8]. Properly designed and analyzed serological surveys mitigate these issues by providing direct measures of population level immunity[9,10].

The optimal design and interpretation of serological surveys depends on many factors, including the antibody class or biomarker measured, the age at which individuals are sampled, the frequency of sampling, the assay used for analysis, etc[1,6]. These features yield different insights into processes associated with the immune landscape (e.g., who is and isn’t protected from infection or disease by pre-existing immunity): cross-sectional serosurveys provide a snapshot of the seropositivity rates and therefore the proportion of individuals protected against a pathogen (which might be used to target vaccination campaigns[1,10]), while longitudinal serosurveys can also yield estimates of seroconversion events between sampling times, and antibody waning rates[11]. Difficulties arise in serosurvey design and analysis as researchers strive to achieve a representative sample of the target population to make generalizations at larger scales, or attain the right temporal sampling density to capture parameters of interest like waning rates[6].

Serosurvey analysis is complicated by the various unobserved and complex immunological and epidemiological processes which generate an individual’s observed biomarker quantity. Statistical and mathematical models designed to interpret observed antibody titers, or biomarkers more generally, range from simple measures of seroconversion or seropositivity (e.g., serocatalytic models [12,13]) to complex models of within- and between-host processes (e.g., hierarchical models of antibody kinetics [14–16]). All of these approaches aim to make useful inferences about exposures and immunity without exhaustively capturing all features of the true data-generating process[8,12,17]. More realistic models describe the link between observed biomarker measurements and latent infection and vaccination states (Fig 1) and can be used to calculate the likelihood of an infection, and estimate antibody kinetics parameters (see Supporting Text 1.2 of Hay et al., 2020 for a full description) [14–16]. These models typically describe key features of the multi-level data-generating process: the population-level processes which govern rates of exposure; the within-host processes which determine immunity and latent antibody kinetics; and the observation process which dictates the relationship between observed and true biomarker quantities (Fig 1). Additionally, hierarchical Bayesian models can help account for the variability from multiple epidemiological processes which can improve our understanding of heterogeneities driving disease dynamics[14,15].

**Fig 1.**
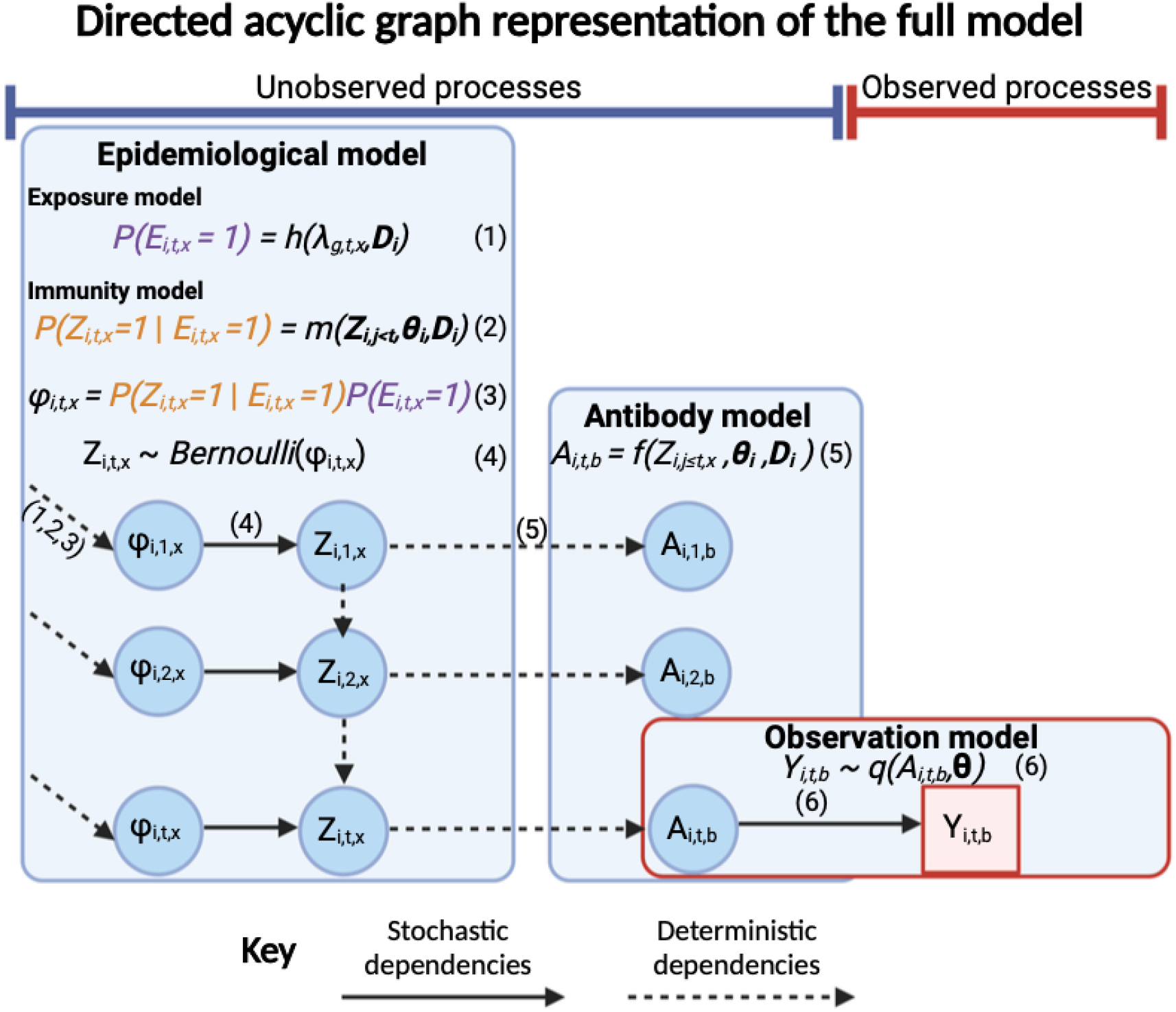
Directed acyclic graph representation of the full *serosim* model. Each model level is shown within a box with stochastic dependencies depicted by a solid arrow and deterministic dependencies by a dashed arrow. Parameters/latent states of interest are depicted within the blue circles while the red square represents the observed state. The unobserved processes level (latent states) contains the epidemiological model (exposure model and immunity model) and the antibody model while the observed processes level contains the observation model. The probability of a successful exposure event *x* for individual *i* at time *t* (*ϕ*_*i,t,x*_) is determined by the user-specified exposure and immunity models (1,2,3) while *Z*_*i,t,x*_ is the binary state indicating whether individual *i* was infected or vaccinated by exposure event *x* at time *t* as determined by a Bernoulli trial (4). The antibody model (5) specifies how the true quantity of biomarker *b* for individual *i* at time *t* (*A*_*i,t,b*_) is generated. Lastly, the observation model (6) specifies how the observed quantity of biomarker *b* for individual *i* at time *t* (*Y*_*i,t,b*_) is generated as a probabilistic function of the true, latent biomarker quantity *A*.

Although new analytical techniques have been developed to aid in the interpretation of serological surveys[9,13,15,16,18], there is a need for common frameworks to simulate observed serological data arising from complex hidden and unobserved epidemiological and immunological processes within different pathogen systems. By simulating synthetic datasets from different complex generative models where the true, full complexity of the data generating process is specified but not observable, such frameworks can be used to assess the performance and trade-offs of new analytical techniques. The *serosim* package allows users to simulate observed biomarker quantities arising from a user-specified set of between- and within-host processes. These simulations provide datasets to evaluate different statistical methods and study designs, allowing users to specify and adjust model inputs to best represent the population and disease system(s) of interest.

We envisage three main use cases for *serosim*:

- Plan serosurvey sampling strategies by simulating various observation processes (varying sample sizes, sampling frequency and assay characteristics) and analyzing the inferences that can be made given the quality of the generated data (e.g., power analyses).
- Uncover the underlying immunological and epidemiological mechanisms responsible for observed biomarkers (e.g., antibody titers) by comparing simulations under different models and parameter values to existing serological survey results.
- Generate synthetic serological data from complex, user-specified generative models to assess the performance and trade-offs of inference models.

Here, we explain the structure of the *serosim* package and its implementation. We then outline the components required for the main function, **runserosim**, explain their purpose and structure, outline useful tools available within *serosim* to aid in their construction and provide a simple example case study. Additionally, we direct users to two additional case study vignettes, available in the package, structured in the same order as the following methods subsections to assist users in constructing the required simulation inputs under different contexts. Lastly, we demonstrate a simple use case for *serosim;* using simulation-recovery experiments to assess the sensitivity and specificity of different biomarker thresholds of seropositivity.

## 1 Methods

### Approach

*Serosim* allows users to define the data-generating process underpinning theoretical serosurvey data with maximum flexibility. To define the different stages of the simulation, we first consider what a generic, full likelihood and prior function would look like to estimate antibody kinetics and epidemiological parameters conditional on a set of observed biomarker quantities in a Bayesian framework (Equation 1, also see [15,19,20]). Each probabilistic term in this equation corresponds to a stochastic event in the simulation (though deterministic models may also be implemented in the same framework). There are a number of indices which must first be described (Table 1). Each individual is denoted *i, t* denotes the discrete time period during which an exposure event or observation could take place and *j* denotes all time periods prior to *t*. Each biomarker which we are concerned with, either as latent quantities or observations are denoted *b*, and may be stimulated by one or more exposure events denoted *x*. Each individual may be associated with a set of demographic information, *D*_*i*_ including a group identifier *g*.

**Table 1.** Names and descriptions of the variables used in the *serosim* package.

| Variable | Name | Description |
| --- | --- | --- |
| **Inputs** |  |  |
| $i$ | <u>I</u>ndividual | Unique individual identifier. |
| $t$ | <u>T</u>ime | Time point at which an individual may be exposed or have a biomarker quantity calculated. |
| $x$ | <u>E</u>xposure event | Identifier for each exposure event which can generate an immunological response in the model (e.g., infection with a SARS-CoV-2 variant or vaccination with a multivalent vaccine). |
| $b$ | <u>B</u>iomarker | Identifier for the biomarker(s) boosted in response to a particular antigen present in the exposure event (e.g., pathogen-specific ELISA IgG titers, microneutralization titers etc.). |
| $g$ | <u>G</u>roup | Group identifier, allowing group-specific force of infection and vaccination parameters (e.g., spatial structure). |
| $D$ | <u>D</u>emography data | Additional demographic data which may impact the infection generation process, the immunity model and/or antibody kinetics (e.g., age-dependent antibody waning rates or vaccination schedule). |
| $\lambda$ | Force of exposure (FOE) | Parameters for the exposure process model, ultimately used to generate a probability of infection or vaccination for a given time period $t$. |
| $\Theta$ | Antibody, immunity and observation model parameters | Parameters governing the within-host process models (e.g., how biomarker quantities change over time-since-infection and how immunity depends on past exposure events). |
| **Outputs** |  |  |
| $\Phi$ | Time varying probability of a successful exposure event | Probability of exposure (determined by the exposure model) multiplied by the probability of a successful exposure event defined as a measurable immunological response (determined by the immunity model). |
| $Z$ | Exposure history array | Array of latent binary states (i.e., 0 or 1) representing the time points and exposure events that an individual has been infected or vaccinated at. |
| $A$ | True biomarker quantity | True, latent biomarker quantity predicted by the antibody kinetics model prior to simulating the observation process. |
| $Y$ | Observed biomarker quantity | Observed biomarker quantity assuming some observation process dictating the distribution of $Y$ as a function of $A$. |

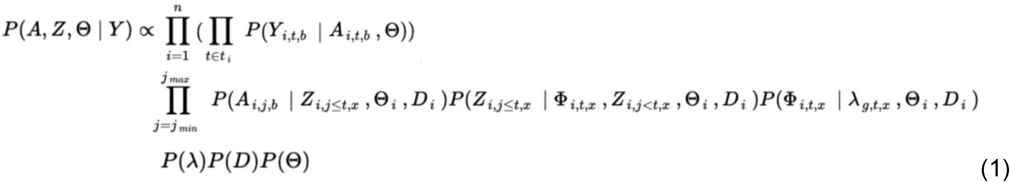

*Serosim* combines the unobserved and observed processes which together contain three different model levels (Fig 1, Table 2). The unobserved processes are composed of the epidemiological model and antibody model. The epidemiological model contains the exposure model *h* which will determine the probability that an individual *i* is exposed in an exposure event *x* and the immunity model *m* which will determine whether that exposure event is successful at generating or boosting biomarker quantities given any relevant factors (e.g., vaccination schedule, titer threshold required for protection, etc.). For the purpose of *serosim*, exposure events *x* are defined as any event which leads to biomarker *b* production, irrespective of any associated disease onset. This definition of exposure allows users to track vaccination, infection and re-infection events simultaneously but separately alongside the biomarkers produced as a result of different exposure events. Biomarkers *b* can represent antibodies binding the entire virus or a specific epitope depending on the user’s preference and assay characteristics. Users will define the biomarker(s) affected by each exposure type within the exposure-to-biomarker map which follows a many-to-many relationship and tells the antibody model *f* which biomarker(s) (e.g., which specific antibody titers) will get boosted and tracked after each exposure event *x*, allowing for simulation of a diverse range of pathogen systems [10,18,21–24] (Fig 2).

**Table 2.**
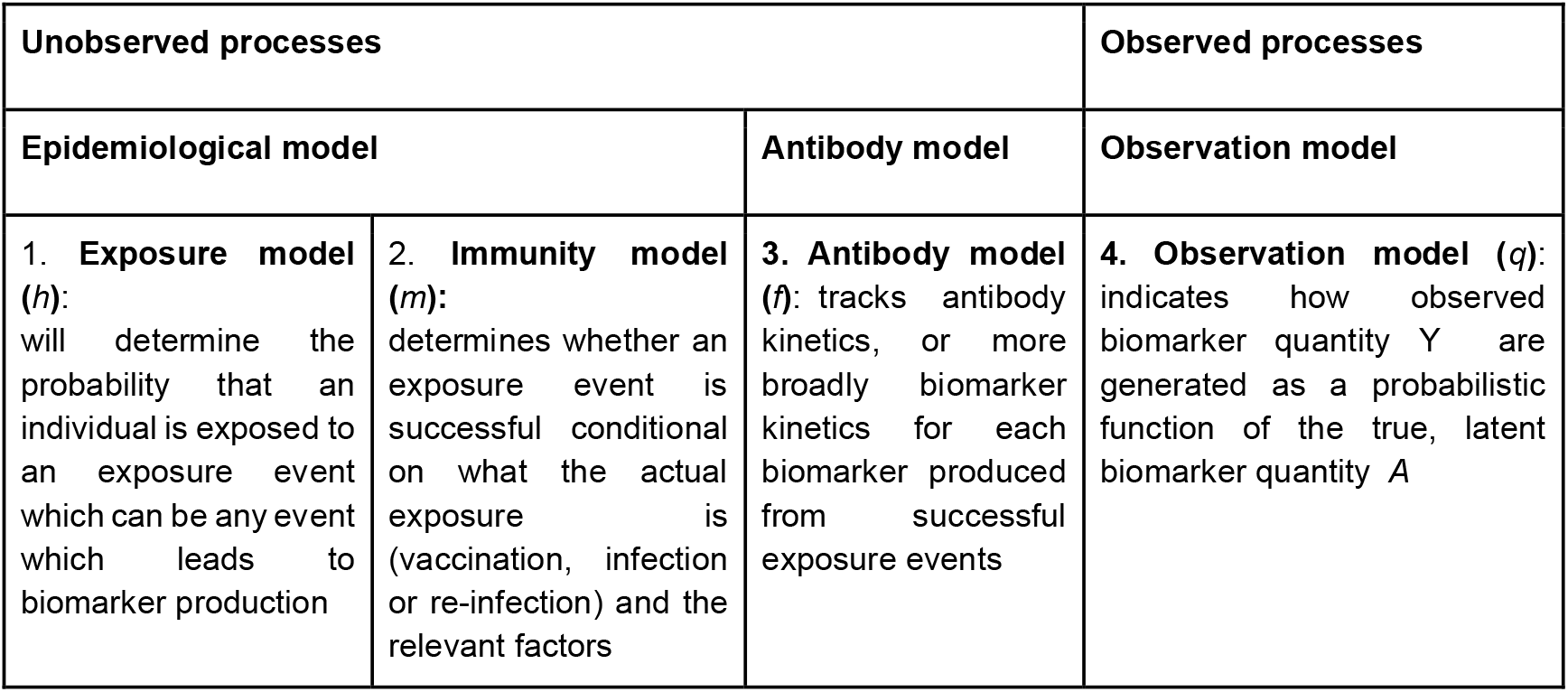
*serosim* framework.

**Fig 2.**
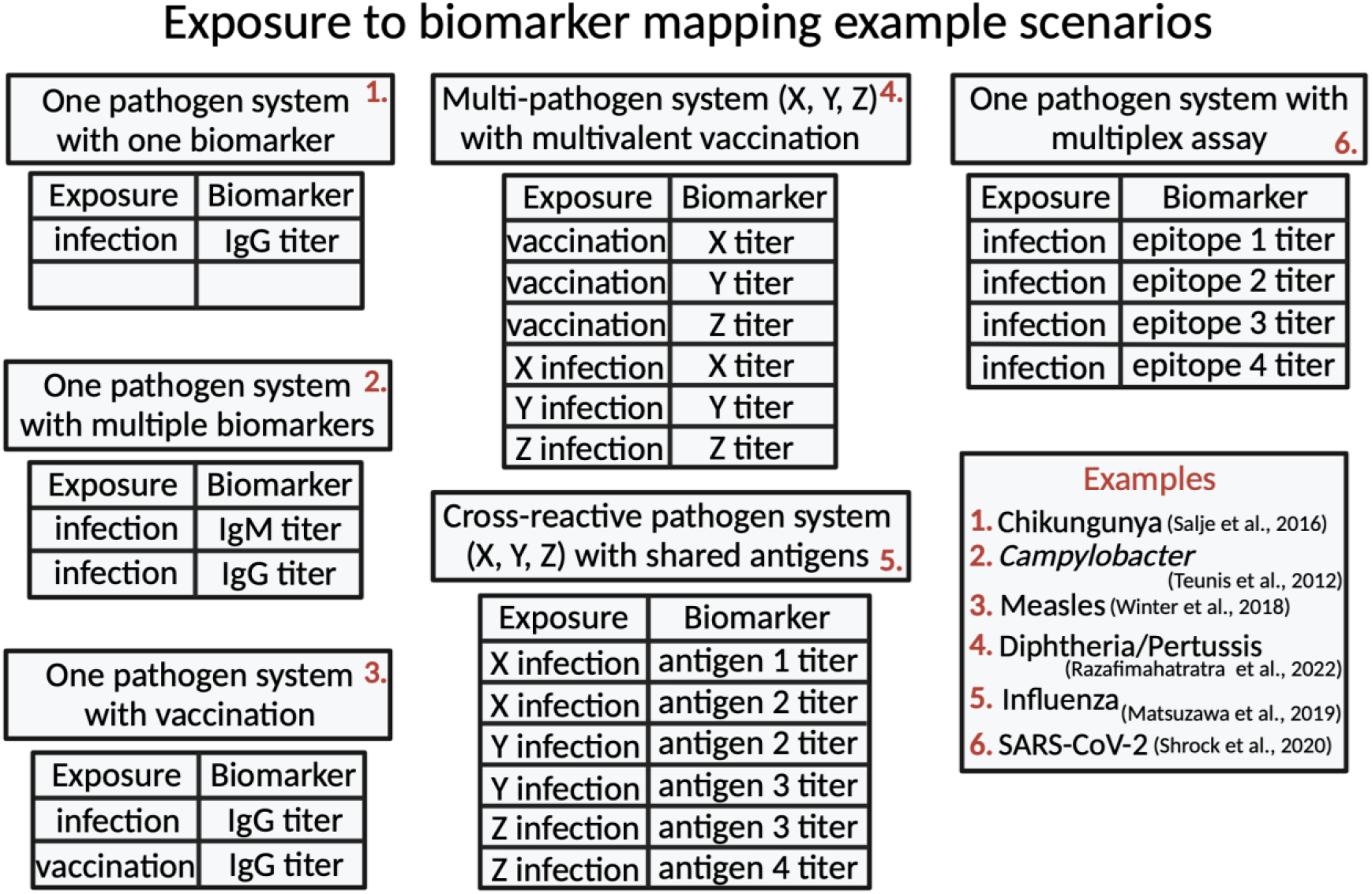
Exposure to biomarker mapping example scenarios. The biomarker map specifies the exposure events *x* and biomarkers *b* of interest for the user’s simulation. Exposure events *x* are defined as any event which leads to biomarker production. Biomarkers *b* can represent antibodies against the entire virus, a specific epitope or a specific antibody class depending on the user’s preference and assay characteristic. Here, we provide various examples of biomarker maps to illustrate the flexibility over the data-generating process provided to the user.

We distinguish between exposure event *x* and biomarker *b* to allow both a single exposure event to boost multiple biomarkers (e.g., antibody titers of multiple antigens following a multivalent vaccine of a single infection) and also to allow the same biomarker to be boosted by multiple exposure events (e.g., both vaccination and natural infection boosting the same biomarker) (Fig 2). This distinction also allows the user to assign different biomarker kinetics parameters (e.g., boosting and waning rates) to different exposure events. Biomarkers can also represent different antibody classes (e.g. IgM and IgG) allowing users to assign different time signatures to biomarkers tracked within the simulation. Users can also incorporate multi-dose vaccines given at different ages by breaking up the vaccine exposure events into multiple exposure events for each vaccine dose. This will allow users to set different age restrictions and antibody kinetics parameters for each dose. Although a majority of *serosim* use cases will track antibodies as biomarkers, we use “biomarker” as a broader term to accommodate newer technologies emerging which don’t measure antibodies directly but rather measure a proportional biomarker[25]. For the purposes of this paper and example vignettes in *serosim*, the term biomarker and antibody are used interchangeably.

The immunity model *m* then determines whether that exposure event *x* is successful conditional on what the actual exposure is (vaccination, infection or re-infection) and any relevant factors, discussed later. The antibody model *f* tracks antibody kinetics, or more broadly biomarker kinetics for each biomarker produced from successful exposure events through time. The exposure, immunity and antibody models make up the complex, unobserved processes responsible for generating an individual’s true biomarker quantity *A* (Fig 1, Table 2).

The second and final level contains the observation model *q* which specifies the serological survey observation process to indicate how observed biomarker quantity *Y* are generated as a probabilistic function of the true, latent biomarker quantity *A* (Fig 1, Table 2). Users can also specify the sampling design (time, frequency and sample size) for the serosurvey and any assay characteristics (sensitivity, specificity and detection limits). Although the underlying, unobserved model inputs can be varied, *serosim* ultimately produces the same output: observed biomarker quantities *Y*.

### Implementation

The *serosim* package is written in the R programming language. Package dependencies are ggplot2[26], data.table[27], tidyverse[28], patchwork[29] and reshape2[30]. All code is publicly available on GitHub (https://github.com/AMenezes97/serosim). For the following sections describing the code, typewriter font refers to function arguments while bold font refers to R functions.

The core of *serosim* is contained within the **runserosim** function, where users specify each of the required model inputs (S1 Table). This function must take a number of default arguments described in S1 Table, but additional optional arguments may also be provided for user-written functions. This gives the user flexibility to tailor their own settings, models and parameters to their population and disease system(s) of interest. For example, if the user wishes to model a fully immunizing pathogen system like measles they can specify an immunity model which only allows individuals with biomarker quantity (e.g. IgG antibody titer) below a specified threshold to become infected and restrict re-infections. If the user wishes to model a pathogen system with multiple infection events and cross-reactivity between different pathogen strains like influenza then they would specify an immunity model where the probability of infection is conditional on an individual’s biomarker quantity (e.g., antibody titer) from any cross-reacting antibodies and allows for multiple infection events. These models would be specified within their respective **runserosim** argument and any additional arguments needed will be passed at the end of **runserosim**. Users are not constrained to data structures of particular dimensions or types, and thus all of the model components can be extended to any desired level of complexity.

The *serosim* package includes tools to help the user generate the necessary inputs for the **runserosim** function, discussed below in their respective sections. Additionally, there are some ready-to-use model functions built into the *serosim* package for convenience (S4-8 Tables). If any of these model functions meet the needs of the user’s disease system and simulation model, the user simply has to specify the desired function as its corresponding argument within the **runserosim** function. These ready-to-use functions also provide a helpful framework if users wish to create their own function or make modifications to an existing function.

The following subsections 1.1-1.7 step through the process of building and specifying the required inputs for a simulation presented in the same order as Figure 3. We have also integrated the README example available within the package within this description.

**Fig 3.**
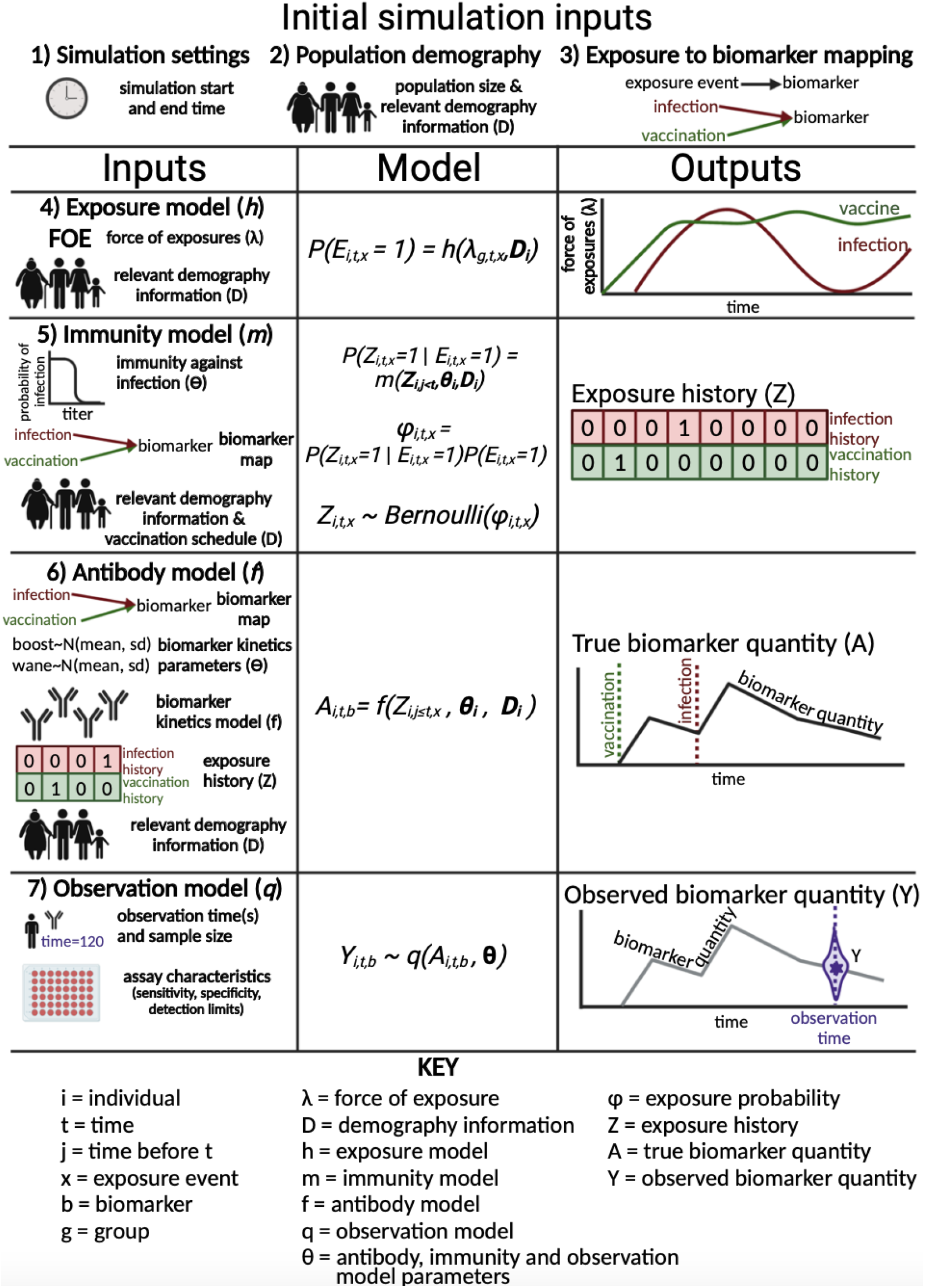
Required inputs, models and subsequent outputs from the main *serosim* function, runserosim. In order to build a simulation with *serosim*, users must follow the 7 steps outlined here and in the methods section to specify the required inputs and models for **runserosim**. Steps 1-3 specify initial simulation inputs while steps 4-7 specify the bulk of the unobserved and observed processes. For steps 4-7, we outline the user-specified inputs in the left column which are used for the user-specified models as depicted by the sampling statements in the middle column. Lastly, the generated outputs produced once the simulation is complete are depicted in the right column.

### 1.1 Simulation settings

Here, the user specifies the start and end time for the simulation. The model will simulate from the initial time point to the final time point in increments of one unless specified otherwise. Note that these are arbitrary time steps selected by the user which will need to be scaled to the right time resolution to match any time-based parameters used in the model (e.g., antibody waning rates). In this example, we simulated a ten year period at the monthly timescale by setting the start and end time points to 1 and 120.

### 1.2 Population demography

Users will specify the population size and have the option of specifying any population demographic elements of interest and relevance for the subsequent models like an individual’s socioeconomic status, nutritional status, sex, group, birth time, etc… via the `demography` tibble [31]. See S1.1 Text for information on how to use the **generate_pop_demography** function to build your `demography` tibble.

For this example, we simulate a population with 100 individuals and we are not interested in tracking any demographic information other than an individual’s birth time. We will use the **generate_pop_demography** function to simulate random birth times and create the `demography` tibble needed for **runserosim**.

### 1.3 Exposure to biomarker mapping

Here, users specify the relationship between exposure event *x* and biomarker(s) *b* (Fig 2). This example assumes one circulating pathogen responsible for a natural infection exposure event (*exposure_ID=ifxn*) and one vaccine exposure event (*exposure_ID=vacc*) both of which will boost the same biomarker (*biomarker_ID=IgG*) (S1 Fig). **runserosim** requires that exposure and biomarker IDs are numeric so we will use the **reformat_biomarker_map** function to create a new version of the biomarker map (S1.2 Text, S1 Fig). Users can go directly to the numeric version if they wish (S1 Fig).

### 1.4 Exposure model

Here, users specify any known information on force of exposure for all exposure events *x* and the first part of the epidemiological model, the exposure model *h*. We distinguish between the probability of exposure (determined by the exposure model) and the probability of an exposure event generating a successful immunological response (determined by the immunity model), so that the latter can take into account important immunological and epidemiological considerations. For example, an individual’s probability of exposure to exposure event *x* which is specified at the population level might be different from their probability of becoming immunized conditional on what the actual exposure event is (vaccination, infection or re-infection) and any relevant factors (e.g. age, number of past vaccinations, current titer level, etc).

The force of the exposure event (`foe_pars`) argument is typically a three-dimensional array indicating the force of exposure for each exposure event in each group (if groups are specified within `demography)` in each time period, though this object can also be another data type for more complex models. Since exposure events can also be vaccination events, we use force of exposure (FOE) rather than force of infection. If there is only one group specified then all individuals will be under the same force of exposure parameter and dimension 1 of `foe_pars` will contain just one row.

Although this package does not include a suite of complex transmission models (e.g., complex Susceptible-Infected-Recovered models), users can incorporate the outputs of their own preferred transmission model within **runserosim**. In `foe_pars`, users can input the force of infection generated from their transmission model into the `exposure_id` dimension associated with natural infection events. Users can also use the optional `exposure_histories_fixed` argument in **runserosim** to include prespecified information on an individual’s exposure history if known (See S2.1 Text and S2 Fig).

The force of exposure (`foe_pars`) array is called by the exposure model function which will determine the probability that an individual is exposed. The exposure model, specified by the `exposure_model` argument within **runserosim**, allows the user to determine which factors are relevant for an individual’s probability of exposure. The exposure model can vary in complexity with simpler versions calculating the probability of exposure by just taking the exposure event’s population level force of exposure into account and with more complex versions providing a functional transformation of the population level force of exposure given an individual’s demographic information specified within `demography`. For example, an individual can be in a group (e.g., a location) where the force of exposure is 0.5 but their socio-economic status is high which might reduce their probability of exposure given that the disease is circulating less in that socio-economic class. See S4 Table for more information on the ready-to-use exposure models included within the *serosim* package. The exposure model is given by:

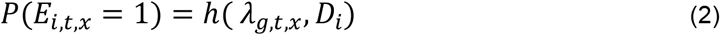

where P(*E*_*i,t,x*_ =1), the probability of an exposure E for individual *i* at time *t* to exposure event *x*, is a function *h* of parameters specified within the **exposure model**, namely *λ*_*g,t,x*_ which represents the force of the exposure (FOE) at time *t* in group *g* from exposure event *x. D*_*i*_ represents relevant demographic information of individual *i*, which may be used to modulate the force of exposure term (e.g., if there are differences in FOE by age).

For simplicity in this example, we assumed a constant force of exposure for exposure event one representing natural infection at *λ*_1,*t*,1_ = 0.01 and for exposure event two representing vaccination at *λ* _1,*t*,2_ = 0.1 for all *t*. Since we did not specify different groups for our individuals within demography, all individuals will automatically be assigned group one within **runserosim**. Therefore, we only need one row for dimension one in `foe_pars`. Dimension two of `foe_pars` represents the total simulation time so there are 120 columns (where the first column is time step one, second column is time step two, etc.) and dimension three is for each exposure event. Similarly, we specified a simple exposure model which calculates the probability of exposure directly from the force of exposure at that time step. In this selected model, the probability of exposure (*1-e*^*-λ*^) depends only on the force of exposure (*λ*) at that time.

### 1.5 Immunity model

The immunity model *m*, specified by the `immunity_model` argument, will determine whether an exposure event is successful in generating an immunological response given any relevant factors specified by the user. The immunity model can vary in complexity with simpler versions assuming all exposures are successful and with more complex versions taking into account what the exposure event is (e.g., a vaccination event or natural exposure to a pathogen), an individual’s past exposure history, current biomarker quantity and any relevant demography data. The immunity model is given by:

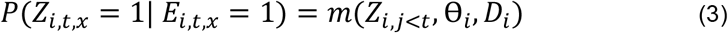

where P(*Z*_*i,t,x*_ =1| *E*_*i,t,x*_ =1), the probability of successful infection or vaccination of exposure event *x* given an exposure, is a function *m* of parameters specified within the immunity model. Z_i,j<t_ represents all successful infection/vaccination events at each timepoint *j* prior to time *t* (i.e., the exposure history), ⊖_*i*_ represents the immunity model parameters and *D*_*i*_ represents any relevant demographic data. Note that P(*Z*_*i,t,x*_ =1| *E*_*i,t,x*_ = 0) = 0 and P(*Z*_*i,t,x*_ = 0| *E*_*i,t,x*_ = 0) =1.

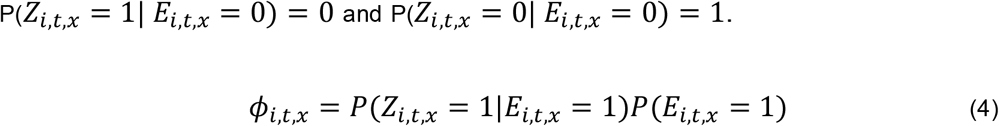

where *ϕ*_*i,t,x*_ is the probability of successful exposure by exposure type *x* for individual *i* at time *t* given the probability of exposure *E*_*i,t,x*_ as determined by the exposure model *h* and the probability of a successful exposure event *Z*_*i,t,x*_ as determined by the immunity model *m*.

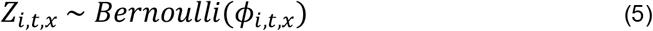

where *Z*_*i,t,x*_ is the binary state indicating whether individual *i* was infected or vaccinated by exposure type *x* at time *t* as determined by a Bernoulli trial.

Users can incorporate immunity from prior infection or vaccination and add limits to vaccine exposure events to prevent vaccinations after a completed series or before an individual is eligible. See S5 Table for more information on the ready-to-use immunity models included within the *serosim* package. Any immunity model parameters *⊖* needed for the immunity model can be specified within `model_pars`.

For this example, we selected a simple immunity model where the probability of a successful exposure event is only conditional on the total number of previous exposure events. With this model, the probability of successful vaccination exposure depends on the number of vaccines received prior to time *t* (*j≤t*) and age at time *t*, while the probability of successful infection is dependent on the number of infections prior to time *t* (*j≤t*). We set both the maximum number of successful vaccination and natural infection events to one and age of vaccine eligibility starting at nine months old.

### 1.6 Antibody model and model parameters

The antibody model *f*, specified by the `antibody_model` argument, is used to track antibody kinetics, or more broadly biomarker kinetics for each biomarker produced following successful exposure events given the biomarker kinetics model parameters specified in `model_pars` and any relevant demography data specified in `demography.` Here, users can specify their preferred model to represent the antibody kinetics process.

The antibody model is formulated to capture the possibility of a probabilistic relationship between exposure history, demography and model parameters with true biomarker quantities. However, in most cases the user will likely assume that biomarker quantity is a deterministic function of exposure history, demography and model parameters. In the deterministic case, latent biomarker quantity *A* for individual *i* at time *t* for biomarker *b* given that individual’s vector of latent infection states ***Z***_***i***_, antibody model parameters *⊖*_*i*_ and relevant demographic information *D*_*i*_ can be generically described by:

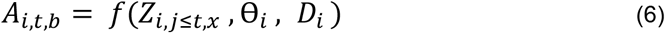

where *A*_*i,t,b*_, the true quantity of biomarker *b* for individual *i* at time *t*, is described by a function *f* conditional on parameters specified within the **antibody model**, including the infection/vaccination history for individual *i* for all exposure events which occurred prior to time *t* (Z_i,j≤t,x_), the antibody model parameters (⊖_*i*_) and any relevant demography data *D*_*i*_.

The model parameters tibble, `model_pars`, specifies any parameters needed for the antibody model. Like the exposure and immunity models, the antibody model can vary in complexity depending on the user’s preferences. For example, users could implement an explicit model of antibody secreting B cells or a biphasic boosting-waning model [32–34].

The last component of the antibody model layer is the **draw_parameters** function which indicates how parameters ⊖ for the antibody model are simulated from `model_pars`. Here, users can either assume a fixed effects or random effects model. A fixed effects model would assume all individuals are governed by the same set of parameters while the random effects model assumes each individual has their own unique set of parameters ⊖ drawn from `model_pars` allowing for individual- and group-level differences in antibody kinetics. See S6 and S7 Tables for more information on the ready-to-use antibody models and **draw_parameters** functions included within the *serosim* package. The antibody model, model parameters tibble (`model_pars`) and **draw_parameters** function must be structured in agreement with each other. The model parameters tibble must contain all of the necessary information needed for the **draw_parameters** function to simulate all required variables in the antibody model.

For this example, we selected a monophasic boosting-waning antibody model where parameters are drawn randomly from a distribution (e.g., see simple hierarchical boosting-waning model with individual heterogeneity for *Salmonella* infection [35]). This antibody model assumes that for each exposure there is a boost and boost waning parameter drawn randomly from a distribution with mean and standard deviation specified within `model_pars` (Table 3).

**Table 3.** `model_pars` **parameters needed for a monophasic boosting-waning antibody model with random effects**

| exposure_ID | biomarker_ID | name | mean | sd | distribution |
| --- | --- | --- | --- | --- | --- |
| ifxn | IgG | boost | 4 | 2 | log-normal |
| ifxn | IgG | wane | 0.0033 | .0005 | log-normal |
| vacc | IgG | boost | 2 | 1 | log-normal |
| vacc | IgG | wane | 0.0016 | .0005 | log-normal |

### 1.7 Observation model

The observation model *q*, specified by `observation_model`, along with `observation_times` are the last required parts of the simulation which specifies the serological survey observation process (Table 2). Here, users can indicate how observed biomarker quantities *Y* are generated as a probabilistic function of the true, latent biomarker quantity *A*. The observation model is given by:

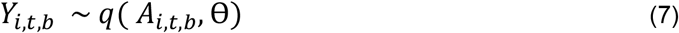

where *Y*_*i,t,b*_, the observed quantity of biomarker *b* for individual *i* at time *t*, is a function *q* specified by the observation model which specifies which distribution to draw the observed quantity where the true biomarker quantity *A*_*i,t,b*_ is the mean and the ⊖ includes the standard deviation as well as any parameters governing assay specificity and sensitivity.

### 1.7a Assay characteristics

To match their assay of choice, users can incorporate assay detection limits like lower bounds, upper bounds, and assay output (discrete vs. continuous titers) within `observation_model` (S8 Table). Users can also incorporate assay sensitivity, specificity and assay noise within the `observation_model`. Noise can be easily added by sampling from a distribution with the true biomarker quantity, *A*, as the mean and the measurement error as the standard deviation such that Y_*i*,t,b_ ∼ N(A_*i*,t,b_, ⊖) where ⊖ is the standard deviation specified within `model_pars` as the “obs_sd” parameter (Table 3). See S8 Table for more information on the ready-to-use observation models included within the *serosim* package. Any model parameters needed for the observation model can be combined into the `model_pars` tibble for convenience. For this example, we selected an observation model which observes the latent biomarker quantity given a continuous assay with added noise (representing assay and sampling variation), no limits of detection and user specified assay sensitivity and specificity. We set the observation standard deviation to 0.25, the assay sensitivity to 85% and the assay specificity to 90%. Therefore, 85% of true positive individuals will have their observed biomarker quantity sampled from a distribution with their true biomarker quantity as the mean and 0.25 as the standard deviation while the other 15% will be classified as a false negative with a reported biomarker quantity of 0. On the other hand, the model will accurately report 90% of true negative individuals’ observed biomarker quantity as 0 with the remaining 10% becoming false positives with an observed biomarker quantity sampled from the range of observed biomarker quantities.

### 1.7b Sampling design

Lastly, users can set the sampling design for the serological survey by indicating the timepoints, individuals and biomarkers to sample from with the `observation_times` input. `observation_times` is a tibble of observation times and biomarkers for each individual. If `observation_times` is not specified within **runserosim** then the simulation will observe quantities of all biomarkers for all individuals at all time steps. For this example, we sample all individuals at the end of the simulation (t=120) for biomarker one (IgG antibody titer).

## 2 Results

Once all arguments have been defined (S1 Table), **runserosim** is ready to go and produce its 6 main outputs (S2 Table). Lastly, users can use available functions to visualize the generated outputs (S3 Table). Here, we have displayed 4 of the **runserosim** outputs for our example simulation (Figs 4 and 5).

**Fig 4.**
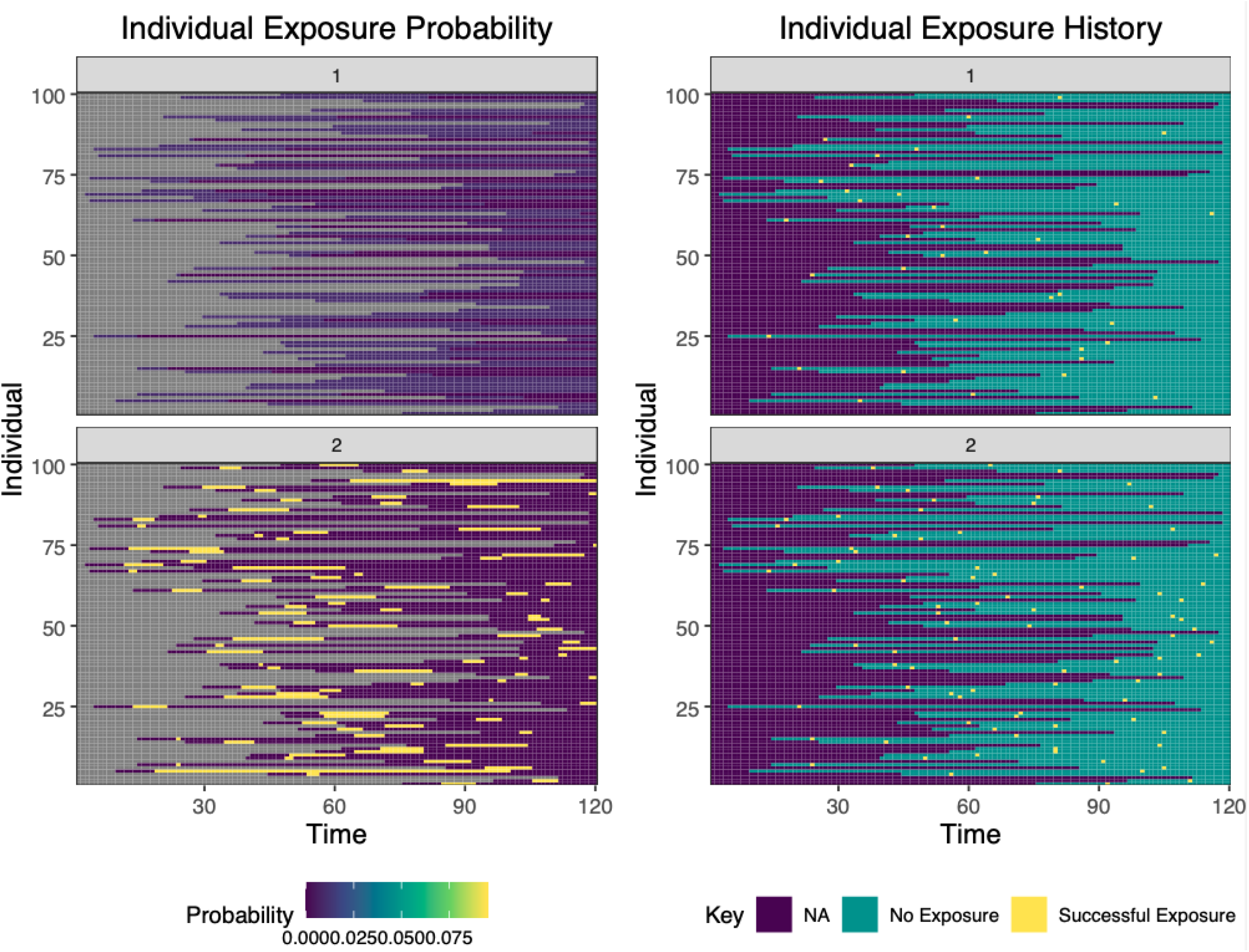
Individual exposure probabilities and individual exposure history plots from a simulation with two exposure types. The left hand plot displays the probability of successful (immunity-boosting) exposure for a simulation with 120 time steps, two exposure events and 100 individuals. The right hand plot displays the exposure histories for the same simulation. Exposure event one (top row) represents an infection event and exposure event two (bottom row) represents a vaccination event. NA indicates that an individual was not available to be exposed in that time period, usually because they were not yet born or entered the study population.

**Fig 5.**
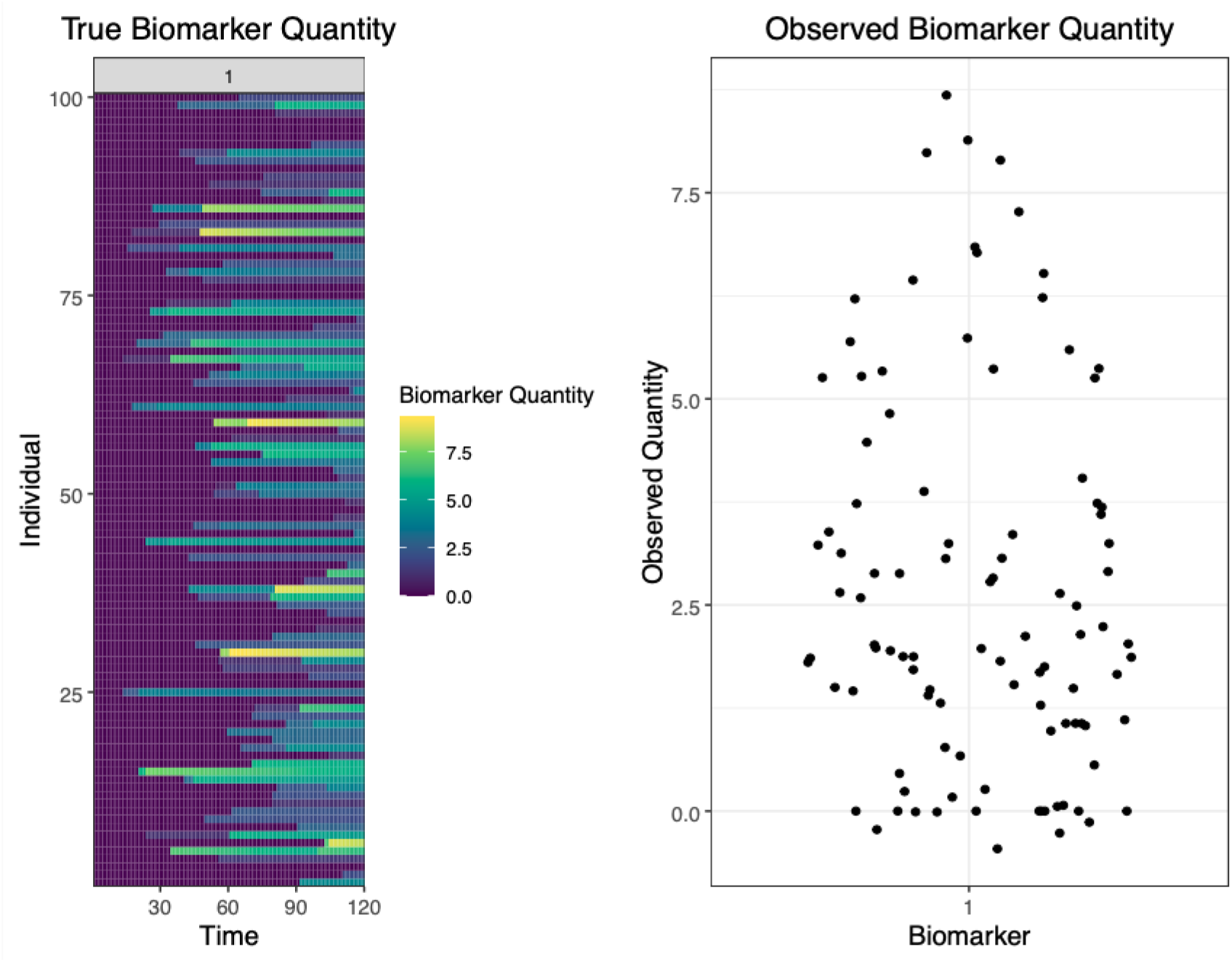
True and observed biomarker quantity plots. The left hand plot displays the true antibody kinetics for a simulation with 120 time steps, one biomarker and 100 individuals. This plot displays all 100 individuals true biomarker quantities for all time steps. The right hand plot displays the observed biomarker quantity at the observation time (t=120) given the specified observation model. In this example, all individuals alive during the endpoint (t=120) had their biomarker quantity, in this case antibody titer, measured with a continuous assay with user-specified noise, sensitivity and specificity. The left hand plot represents the unobserved process level within *serosim* generated by the exposure, immunity and antibody models while the right hand plot represents the observed data generated by the observation model (Fig 1). The true antibody kinetics for each individual (left hand plot) is not known in real world settings where researchers only have cross-sectional antibody titers (right hand plot).

### 2.1 Case studies

The *serosim* package contains two example case study vignettes to illustrate how to use *serosim* and how to structure the required inputs for **runserosim**. Case study 1 provides an example of a longitudinal singular biomarker serological survey simulation structured around measles, a one-pathogen system with vaccination, but also applicable to other vaccine preventable diseases. Case study 2 provides an example of a cross-sectional multi-biomarker serological survey structured around diphtheria and pertussis, a two-pathogen system with bivalent vaccination, but also applicable to multi-pathogen systems with multivalent vaccines. Figure 6 provides run times for simulations of varying complexity by adjusting the number of individuals for the README example and 2 case studies. *serosim* is not limited to either of these scenarios and we hope it will have applications in other systems including wildlife pathogens[20,36].

**Fig 6.**
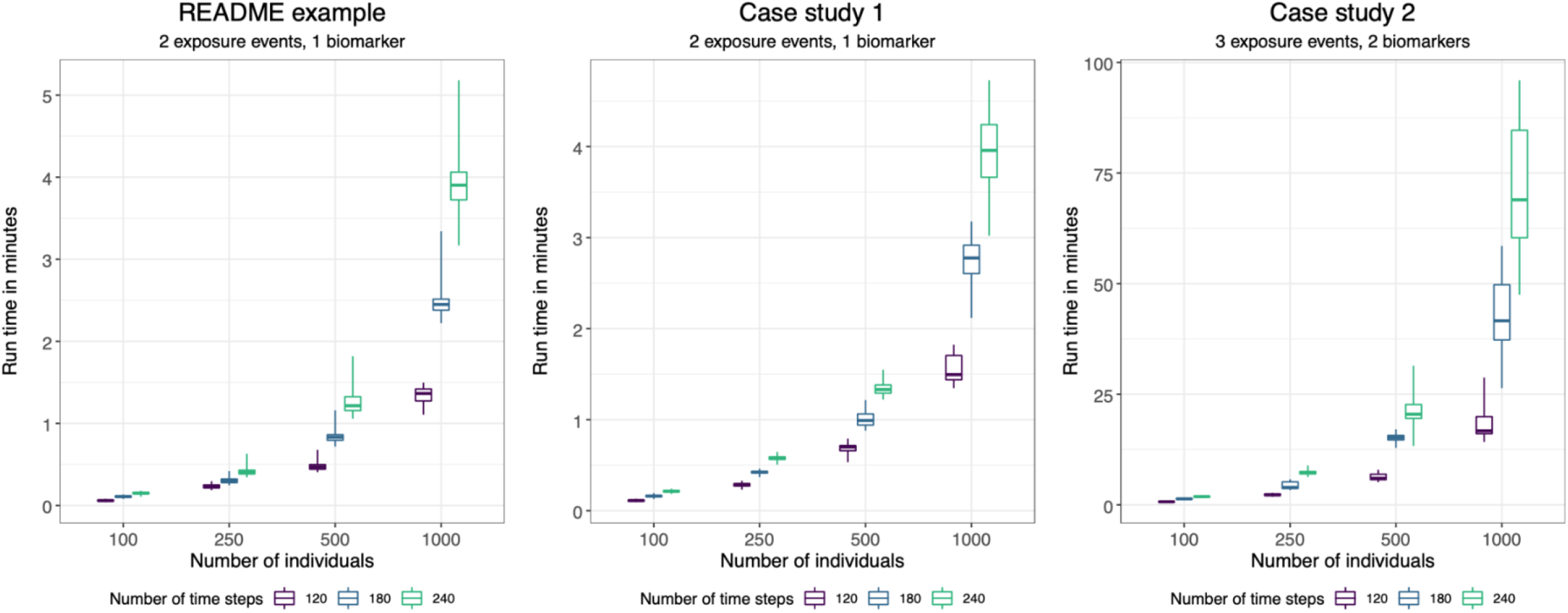
runserosim run times for simulations of varying complexities. We ran the **runserosim** function 100 times and report the mean and range of run times under various simulation settings (number of individuals and time steps). All three of these example cases are included in *serosim*. As you move to the right, model complexity increases so that case study 2 uses more computationally complex exposure, immunity, antibody, and observation models than the README example. Case study 1 is similar to the README in the number of exposure events and biomarkers but uses more realistic and complex exposure, immunity, antibody and observation models. The increase in run time between case study 1 and case study 2 is due to a computationally complex exposure model which modifies each individual’s force of exposure based on their age and nutritional status.

### 2.2 Sensitivity and specificity for varying thresholds of seropositivity

Lastly, we briefly explore the sensitivity and specificity of various seropositivity thresholds within the context of case study one. The seropositivity threshold is commonly used to separate seropositive and therefore immune individuals from seronegative individuals who might be susceptible. This threshold is typically set by the assay manufacturer but can also be adjusted by the user to conduct more specific and sensitive analyses.

Since each individual’s exposure history is known in *serosim*, we can assess the sensitivity and specificity tradeoffs between assigning different thresholds for seropositivity. We ran case study one (based on measles) which simulates 100 individuals across 120 time steps 100 times and examined the sensitivity and specificity of varying measles thresholds of seropositivity ranging from 100 milli-international units per milliliter (mIU/mL) (the lower limit of detection of the ELISA assay with which case study one was structured around) to 350 mIU/mL (Fig 7). Sensitivity remains at 99.56% while specificity increases from 83.68% at 100 mIU/mL to 99.95% at 350 mIU/mL (Fig 7). In this scenario, sensitivity remains high because of the large quantity of true positives and low number of false negatives as a result of the 99.6% reported assay sensitivity, the fact that there isn’t much noise attributed to the antibody measurement step and ultimately that biomarker boosts due to vaccination and natural infection is high. In a system with more noise and lower assay sensitivity or exposure events with smaller biomarker boosts or faster waning rates, we would expect more variation in the sensitivity of different seropositivity thresholds. This example demonstrates how *serosim* can be used to assess the accuracy of classification criteria based on limited observations when the true latent state of the system is known and to explore the impact of epidemiological context and within-host processes on diagnostic performance.

**Fig 7.**
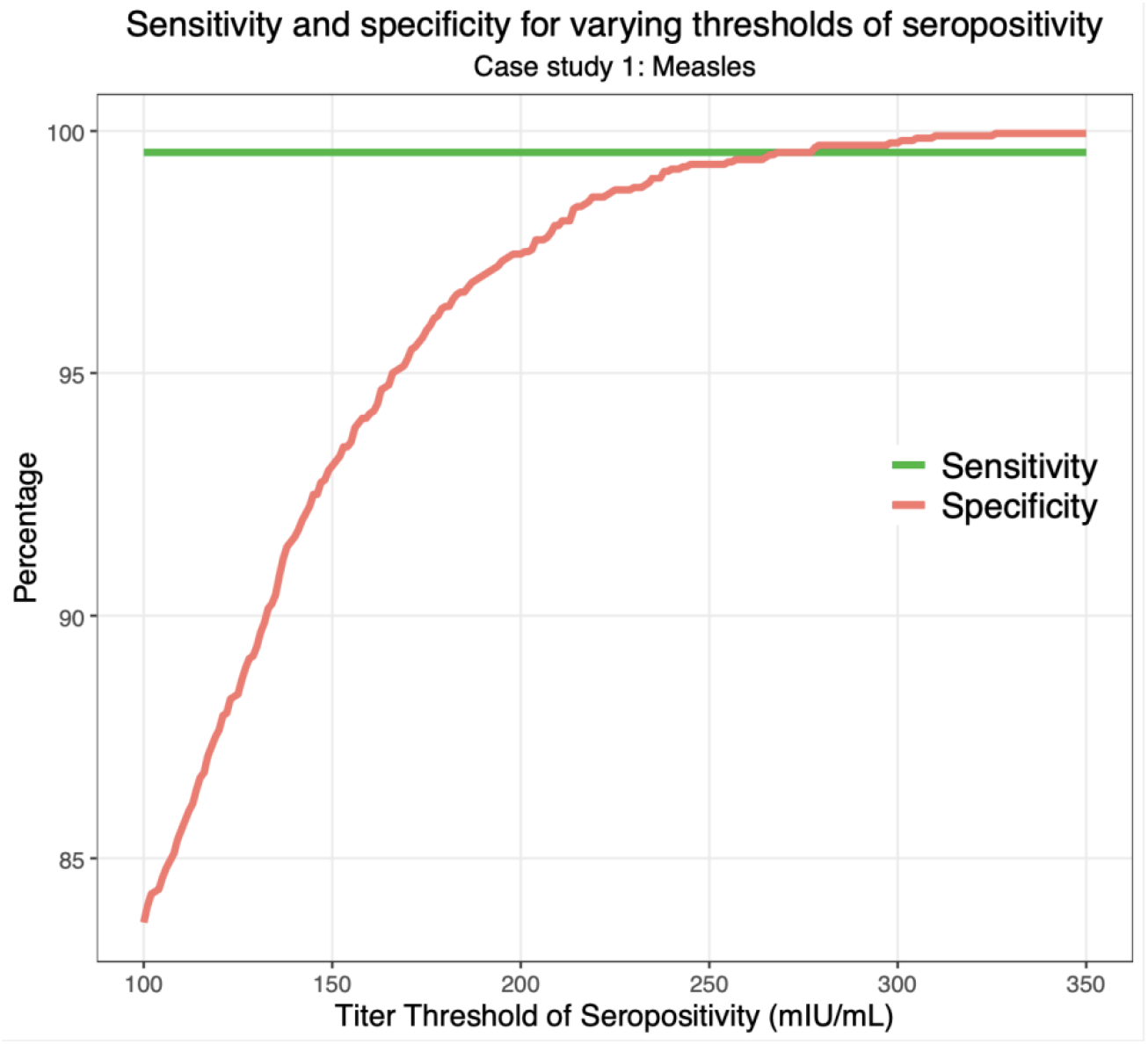
Sensitivity and specificity for varying thresholds of seropositivity in simulation-recovery experiments. We simulated case study one 100 times and stored individual exposure history and observed biomarker quantities. We then calculated the number of true positives, true negatives, false positives and false negatives for identifying infections using various titer thresholds for seropositivity ranging from 100 mIU/mL to 350 mIU/mL. Here, we plotted the sensitivity and specificity achieved at each of those titer thresholds.

## Conclusion, limitations and future directions

The *serosim* package was designed to simulate serological data under different survey designs and underlying immunoepidemiological processes while also providing a systematic framework for validating inference methods. Many epidemiological studies using serological data rely on pre-defined definitions of seroconversion, seropositivity or antibody kinetics to draw inferences about the underlying epidemiology of a pathogen[37]. However, such studies often use real-world datasets without first considering the biases that may arise when the fitted model differs substantially from the true data-generating process. Simulation studies are useful for identifying the limitations, biases and accuracies arising from sample variability, misspecified models or neglected variables[16,38–40]. *serosim* is a generalized approach to this problem, allowing researchers to simulate serological data representing a disease system of interest by explicitly specifying the within-host processes, patterns of infection and vaccination, population demography and other factors which determine observed biomarker quantities. By generating simulations within a single framework, we anticipate better generalization and comparison of methods between pathogen systems and serologic assays[1,14,41].

A key use case for *serosim* is for researchers and public health officials to improve serological survey design. We envision that *serosim* can be used to simulate datasets with different sample sizes, spatial distribution of samples, and sampling frequencies, providing a tool for power calculations. There are a number of existing frameworks for evaluating survey designs based on seroprevalence estimates, taking into account both the contribution of imperfect sensitivity, specificity and model variability to parameter estimation uncertainty[16,42]. However, few tools explicitly consider the full complexity of the underlying biology and epidemiology which determine an observed sample of antibody titers[40]. By separating each step of this generative process into distinct modules, *serosim* requires users to account for sources of measurement error, variation and bias that cause deviation from the underlying parameters of interest. With this generative model in place, inference models (e.g., estimating force of infection using a serocatalytic model [12] or estimating time-since-infection using antibody waning estimates[35]) can be applied to simulated datasets of different sizes and sampling designs to assess their power in recovering ground-truth parameters. These so-called “simulation-recovery” experiments are especially useful as the model being fitted will be, by definition, a misspecified version of the true generative model (with parallels to statistical model misspecification [43]). Designing studies using these simulated datasets also provides interim data with which researchers can develop their inference methods as data collection progresses.

Although we have structured the *serosim* model and code in a flexible, generalized framework, there are limitations to its ease of application to some systems. Some parts of the generative process are very poorly understood for some pathogens (e.g., correlates of protection for the immunity model may be complex[44]; the relationship between time-since-infection and multiple biomarker quantities may be highly variable[45]), and the relationships between exposure and measurements of some modern multiplex serological assays are not yet precisely characterized[25]. We have included a range of ready-to-use functions to encompass various epidemiological and immunological contexts here (S4-8 Tables), however, this is not an exhaustive list as we encourage users to construct new model inputs to match their desired system (e.g., constructing their own antibody kinetics model). There is also a trade-off between framework flexibility and computational speed. Structuring *serosim* as distinct modules places responsibility on the user to ensure that custom code is fast enough to not substantially bottleneck the simulation, particularly for systems with a large number of exposure types, biomarkers, or individuals. However, given that *serosim* is not intended for integration in multi-iteration code (e.g., nested within a Markov chain Monte Carlo algorithm), speed is unlikely to be an issue for most use cases.

Part of the motivation behind *serosim* is to support the rise in development of complex models relying on quantitative antibody measurements as opposed to simple binary serostatus data. These models explicitly describe the relationship between antibody titer and time-since-infection, often estimated using longitudinal data, and then use this relationship to back-calculate seroincidence from cross-sectional data [13,14,35,46–48]. Although underpinned by similar principles, such approaches are usually tailored to a particular pathogen system. By encouraging researchers to be explicit about the generative process common to many settings, we hope to facilitate the sharing and standardization of these more advanced sero-epidemiologic methods.

## Data Availability

All code, tutorials and documentation are made freely available under the GNU General Public License at https://github.com/AMenezes97/serosim. Commit 65c7163 was used at the time of submission.

## Acknowledgments

Figures 1-3 and S1 Fig were created with BioRender.com.

## Financial support

CJEM was supported by a Bill and Melinda Gates Foundation grant (INV-016091). CJEM gratefully acknowledges financial support from the Schmidt DataX Fund at Princeton University made possible through a major gift from the Schmidt Futures Foundation. JAH is funded by a Wellcome Trust Early Career Award (225001/Z/22/Z).

## Conflict of interest

The authors have no conflicts of interest to report.

## Notes

### Competing Interest Statement

The authors have declared no competing interest.

## References

1. Metcalf CJE, Farrar J, Cutts FT, Basta NE, Graham AL, Lessler J, et al. Use of serological surveys to generate key insights into the changing global landscape of infectious disease. Lancet. 2016;388: 728–730.

2. Takahashi S, Metcalf CJE, Ferrari MJ, Moss WJ, Truelove SA, Tatem AJ, et al. Reduced vaccination and the risk of measles and other childhood infections post-Ebola. Science. 2015;347: 1240–1242.

3. Bjørnstad ON, Finkenstädt BF, Grenfell BT. Dynamics of Measles Epidemics: Estimating Scaling of Transmission Rates Using a Time Series SIR Model. Ecol Monogr. 2002;72: 169–184.

4. Lessler J, Metcalf CJE, Grais RF, Luquero FJ, Cummings DAT, Grenfell BT. Measuring the performance of vaccination programs using cross-sectional surveys: a likelihood framework and retrospective analysis. PLoS Med. 2011;8: e1001110.

5. Cutts FT, Izurieta HS, Rhoda DA. Measuring coverage in MNCH: design, implementation, and interpretation challenges associated with tracking vaccination coverage using household surveys. PLoS Med. 2013;10: e1001404.

6. Winter AK, Martinez ME, Cutts FT, Moss WJ, Ferrari MJ, McKee A, et al. Benefits and Challenges in Using Seroprevalence Data to Inform Models for Measles and Rubella Elimination. J Infect Dis. 2018;218: 355–364.

7. Murray CJL, Shengelia B, Gupta N, Moussavi S, Tandon A, Thieren M. Validity of reported vaccination coverage in 45 countries. Lancet. 2003;362: 1022–1027.

8. Clapham H, Hay J, Routledge I, Takahashi S, Choisy M, Cummings D, et al. Seroepidemiologic Study Designs for Determining SARS-COV-2 Transmission and Immunity. Emerg Infect Dis. 2020;26: 1978–1986.

9. Cucunubá ZM, Nouvellet P, Conteh L, Vera MJ, Angulo VM, Dib JC, et al. Modelling historical changes in the force-of-infection of Chagas disease to inform control and elimination programmes: application in Colombia. BMJ Glob Health. 2017;2: e000345.

10. Winter AK, Wesolowski AP, Mensah KJ, Ramamonjiharisoa MB, Randriamanantena AH, Razafindratsimandresy R, et al. Revealing Measles Outbreak Risk With a Nested Immunoglobulin G Serosurvey in Madagascar. Am J Epidemiol. 2018;187: 2219–2226.

11. Amanna IJ, Carlson NE, Slifka MK. Duration of humoral immunity to common viral and vaccine antigens. N Engl J Med. 2007;357: 1903–1915.

12. Hens N, Aerts M, Faes C, Shkedy Z, Lejeune O, Van Damme P, et al. Seventy-five years of estimating the force of infection from current status data. Epidemiol Infect. 2010;138: 802–812.

13. Salje H, Cummings DAT, Rodriguez-Barraquer I, Katzelnick LC, Lessler J, Klungthong C, et al. Reconstruction of antibody dynamics and infection histories to evaluate dengue risk. Nature. 2018;557: 719–723.

14. Pepin KM, Kay SL, Golas BD, Shriner SS, Gilbert AT, Miller RS, et al. Inferring infection hazard in wildlife populations by linking data across individual and population scales. Ecol Lett. 2017;20: 275–292.

15. Hay JA, Minter A, Ainslie KEC, Lessler J, Yang B, Cummings DAT, et al. An open source tool to infer epidemiological and immunological dynamics from serological data: serosolver. PLoS Comput Biol. 2020;16: e1007840.

16. Larremore DB, Fosdick BK, Bubar KM, Zhang S, Kissler SM, Metcalf CJE, et al. Estimating SARS-CoV-2 seroprevalence and epidemiological parameters with uncertainty from serological surveys. Elife. 2021;10. doi:10.7554/eLife.64206

17. Pothin E, Ferguson NM, Drakeley CJ, Ghani AC. Estimating malaria transmission intensity from Plasmodium falciparum serological data using antibody density models. Malar J. 2016;15: 79.

18. Salje H, Cauchemez S, Alera MT, Rodriguez-Barraquer I, Thaisomboonsuk B, Srikiatkhachorn A, et al. Reconstruction of 60 Years of Chikungunya Epidemiology in the Philippines Demonstrates Episodic and Focal Transmission. J Infect Dis. 2016;213: 604–610.

19. Rydevik G, Innocent GT, Marion G, Davidson RS, White PCL, Billinis C, et al. Using Combined Diagnostic Test Results to Hindcast Trends of Infection from Cross-Sectional Data. PLoS Comput Biol. 2016;12: e1004901.

20. Borremans B, Hens N, Beutels P, Leirs H, Reijniers J. Estimating Time of Infection Using Prior Serological and Individual Information Can Greatly Improve Incidence Estimation of Human and Wildlife Infections. PLoS Comput Biol. 2016;12: e1004882.

21. Teunis PFM, van Eijkeren JCH, Ang CW, van Duynhoven YTHP, Simonsen JB, Strid MA, et al. Biomarker dynamics: estimating infection rates from serological data. Stat Med. 2012;31: 2240–2248.

22. Matsuzawa Y, Iwatsuki-Horimoto K, Nishimoto Y, Abe Y, Fukuyama S, Hamabata T, et al. Antigenic Change in Human Influenza A(H2N2) Viruses Detected by Using Human Plasma from Aged and Younger Adult Individuals. Viruses. 2019;11. doi:10.3390/v11110978

23. Shrock E, Fujimura E, Kula T, Timms RT, Lee I-H, Leng Y, et al. Viral epitope profiling of COVID-19 patients reveals cross-reactivity and correlates of severity. Science. 2020;370. doi:10.1126/science.abd4250

24. Razafimahatratra SL, Menezes A, Wesolowski A, Rafetrarivony L, Cauchemez S, Razafindratsimandresy R, et al. Leveraging serology to titrate immunisation programme functionality for diphtheria in Madagascar. Epidemiology & Infection. 2022;150. doi:10.1017/S0950268822000097

25. Mina MJ, Kula T, Leng Y, Li M, de Vries RD, Knip M, et al. Measles virus infection diminishes preexisting antibodies that offer protection from other pathogens. Science. 2019;366: 599–606.

26. Wickham H, Navarro D, Pedersen TL. ggplot2: Elegant Graphics for Data Analysis; 2009.

27. Dowle, Srinivasan, Short. data. table: Extension of “data. frame.” R package version.

28. Wickham H, Averick M, Bryan J, Chang W, McGowan L, François R, et al. Welcome to the tidyverse. J Open Source Softw. 2019;4: 1686.

29. Pedersen TL. Package “patchwork.” R package http://CRANR9J8MH6t3BYCPXMFLa35R7aUJV9i4siPhX. 2019. Available: ftp://ftp.onet.pl/pub/mirrors/CRAN/web/packages/patchwork/patchwork.pdf

30. Wickham H. reshape2: flexibly reshape data: a reboot of the reshape package. R package version.

31. Wickham KMA. tibble: Simple Data Frames. 2022. Available: https://CRAN.R-project.org/package=tibble

32. Le D, Miller JD, Ganusov VV. Mathematical modeling provides kinetic details of the human immune response to vaccination. Front Cell Infect Microbiol. 2014;4: 177.

33. White MT, Griffin JT, Akpogheneta O, Conway DJ, Koram KA, Riley EM, et al. Dynamics of the antibody response to Plasmodium falciparum infection in African children. J Infect Dis. 2014;210: 1115–1122.

34. Andraud M, Lejeune O, Musoro JZ, Ogunjimi B, Beutels P, Hens N. Living on three time scales: the dynamics of plasma cell and antibody populations illustrated for hepatitis a virus. PLoS Comput Biol. 2012;8: e1002418.

35. Simonsen J, Mølbak K, Falkenhorst G, Krogfelt KA, Linneberg A, Teunis PFM. Estimation of incidences of infectious diseases based on antibody measurements. Stat Med. 2009;28: 1882–1895.

36. Prager KC, Buhnerkempe MG, Greig DJ, Orr AJ, Jensen ED, Gomez F, et al. Linking longitudinal and cross-sectional biomarker data to understand host-pathogen dynamics: Leptospira in California sea lions (Zalophus californianus) as a case study. PLoS Negl Trop Dis. 2020;14: e0008407.

37. Hens N, Shkedy Z, Aerts M, Faes C, Van Damme P, Beutels P. Modeling Infectious Disease Parameters Based on Serological and Social Contact Data: A Modern Statistical Perspective. Springer; 2014.

38. Gamble A, Garnier R, Chambert T, Gimenez O, Boulinier T. Next-generation serology: integrating cross-sectional and capture-recapture approaches to infer disease dynamics. Ecology. 2020;101: e02923.

39. Wu KM, Riley S. Simulation-guided design of serological surveys of the cumulative incidence of influenza infection. BMC Infect Dis. 2014;14: 505.

40. Vinh DN, Boni MF. Statistical identifiability and sample size calculations for serial seroepidemiology. Epidemics. 2015;12: 30–39.

41. Mina MJ, Metcalf CJE, McDermott AB, Douek DC, Farrar J, Grenfell BT. A global lmmunological observatory to meet a time of pandemics. Elife. 2020;9: 1–5.

42. Blaizot S, Herzog SA, Abrams S, Theeten H, Litzroth A, Hens N. Sample size calculation for estimating key epidemiological parameters using serological data and mathematical modelling. BMC Med Res Methodol. 2019;19: 51.

43. Sivula T, Magnusson M, Matamoros AA, Vehtari A. Uncertainty in Bayesian Leave-One-Out Cross-Validation Based Model Comparison. 2020 [cited 24 Nov 2022]. doi:10.48550/arXiv.2008.10296

44. Feng S, Phillips DJ, White T, Sayal H, Aley PK, Bibi S, et al. Correlates of protection against symptomatic and asymptomatic SARS-CoV-2 infection. Nat Med. 2021;27: 2032–2040.

45. Helb DA, Tetteh KKA, Felgner PL, Skinner J, Hubbard A, Arinaitwe E, et al. Novel serologic biomarkers provide accurate estimates of recent Plasmodium falciparum exposure for individuals and communities. Proc Natl Acad Sci U S A. 2015;112: E4438–47.

46. Pelleau S, Woudenberg T, Rosado J, Donnadieu F, Garcia L, Obadia T, et al. Kinetics of the Severe Acute Respiratory Syndrome Coronavirus 2 Antibody Response and Serological Estimation of Time Since Infection. J Infect Dis. 2021;224: 1489–1499.

47. Kucharski AJ, Lessler J, Cummings DAT, Riley S. Timescales of influenza A/H3N2 antibody dynamics. PLoS Biol. 2018;16: e2004974.

48. Tsang TK, Perera RAPM, Fang VJ, Wong JY, Shiu EY, So HC, et al. Reconstructing antibody dynamics to estimate the risk of influenza virus infection. Nat Commun. 2022;13: 1557.

