## Supplement for "*serosim*: an R package for simulating serological survey data arising from vaccination, epidemiological and antibody kinetics processes"

### Supporting information

#### Supplementary Text

**S1 Text.** Helpful functions used to generate inputs for **runserosim**
**S2 Text.** Optional **runserosim** arguments

#### Supplementary Figures

**S1 Fig. Example `biomarker_map` before and after reformatting.**
**S2 Fig. Example `exposure_histories` array**

#### Supplementary Tables

**S1 Table.** Names and descriptions of the main arguments required in the runserosim function.
**S2 Table.** Description of `runserosim` outputs.
**S3 Table.** Description of functions to plot `runserosim` outputs.
**S4 Table.** Names and descriptions of the ready-to-use exposure models included in serosim.
**S5 Table.** Names and descriptions of the ready-to-use immunity models included in serosim.
**S6 Table.** Names and descriptions of the ready-to-use antibody models and model_pars files included in serosim.
**S7 Table.** Names and descriptions of the ready-to-use draw_parameters functions included in serosim.
**S8 Table.** Names and descriptions of the ready-to-use observation models included in serosim.

### Supplementary Text

#### S1 Text. Helpful functions used to generate inputs for runserosim

**S1.1 Text. generate_pop_demography:** This function helps users build their demography tibble with birth times, removal times, and specific demographic elements of interest. This function calls both the **simulate_birth_times** and **simulate_removal_times** functions which simulate random birth times and removal times for each individual. See the **generate_pop_demography** help file for more information on how to specify the limits and probabilities of birth and removal times. With the `aux` in **generate_pop_demography**, users can specify other demographic elements of interest by passing through a list of the column name, options and distributions for each option type. The returned demography tibble can be modified post-hoc to use user-specified distributions and values.

**S1.2 Text. reformat_biomarker_map**: This function will reformat the `biomarker_map` or `model_pars` objects so that exposure_ID and biomarker_ID are either both numeric (if passed as characters) or characters (if passed as numeric) (See S1 Fig). **runserosim** requires that both exposure_ID and biomarker_ID are numeric entries.

**S1.3 Text. plot_biomarker_mediated_protection:** If the user has specified an immunity model with biomarker mediated-protection, they can use this function to produce a plot of the probability of infection given an individual's biomarker quantity at exposure conditional on `biomarker_prot_midpoint` and `biomarker_prot_width` which are specified within `model_pars`. Ready-to-use immunity models which incorporate biomarker mediated protection are `immunity_model_ifxn_biomarker_prot` and `immunity_model_vacc_ifxn_biomarker_prot` (S5 Table). Users can use this function to determine the best values for these two inputs within `model_pars`.

**S1.4 Text. plot_biomarker_dependent_boosting:** If the user has specified a function to draw parameters with biomarker quantity dependent boosting effects, they can use this function to produce a plot displaying the proportion of full boost received at each starting biomarker quantity given the biomarker quantity ceiling threshold and the biomarker quantity ceiling gradient which are specified within `model_pars`. Ready-to-use draw_parameters functions which incorporate biomarker dependent boosting are `draw_parameters_fixed_fx_biomarker_dep` and `draw_parameters_random_fx_biomarker_dep` (S7 Table). Users can use this function to determine the best values for these two inputs within `model_pars`.

#### S2 Text. Optional runserosim arguments

**S2.1 Text. `exposure_histories_fixed`:** The exposure histories array, specified by `exposure_histories`, is a 3-dimensional array indicating the exposure history (1 = exposed; 0 = not exposed) for each individual (dimension 1) at each time (dimension 2) for each exposure event (aka exposure_ID) (dimension 3) (S2 Fig). This array normally contains NA's but can also include pre-specified information if exposure histories are known which can be passed to

**runserosim** with `exposure_histories_fixed`. Wherever an entry is provided, **runserosim** assumes that exposure time and type are known and fixed (e.g., specifying known vaccination times or specifying known infections from a transmission model). If the entry is left as NA, then **runserosim** simulates an exposure event for that entry using the specified models.

**S2.2 Text.** `verbose`: If an integer is specified then a progress update will be printed once the simulation reaches that individual and every multiple thereafter.

**S2.3 Text.** `attempt_precomputation`: If TRUE, the simulation attempts to perform as much pre-computation as possible for the exposure model to speed up the main simulation code. If FALSE, the simulation skips this step.

**S2.4 Text.** Model specific arguments: The user may need to specify additional arguments required for the ready-to-use functions that they select. Refer to the function help file for more information on required arguments.

#### Supplementary Figures

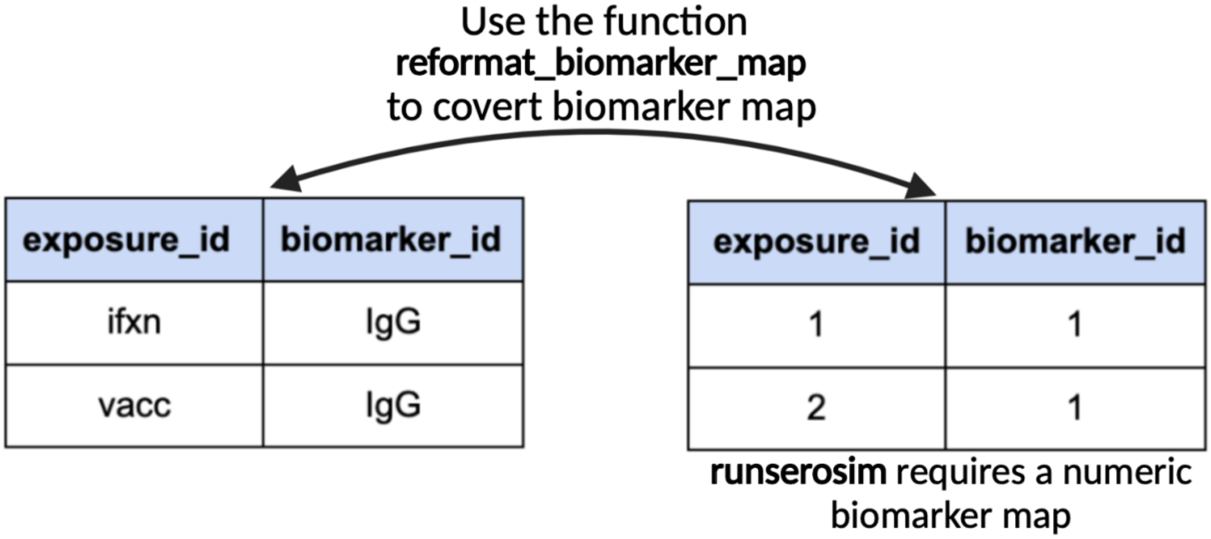


**S1 Fig. Example `biomarker_map` before and after reformatting.** The **runserosim** function requires a numeric biomarker map input as seen on the right. In this example, we have two exposure events (*exposure_id = ifxn and vacc*) and we are interested in tracking one biomarker (*biomarker_id=IgG*) produced by both exposure events.

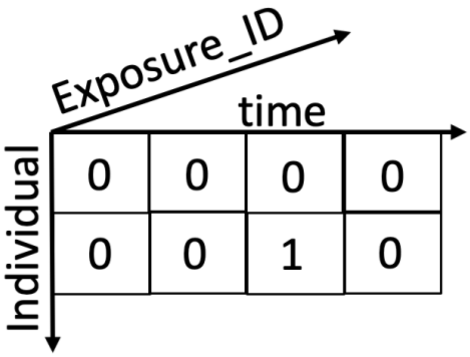


**S2 Fig. Example `exposure_histories` array.** The `exposure_histories` argument is a 3-dimensional array indicating the individual (dimension 1) at each time (dimension 2) for each exposure event (dimension 3). Here, individual 1 never had a successful exposure while individual 2 was exposed at time 3. (See S2.1 Text)

**S1 Table. Names and descriptions of the main arguments required in the runserosim function.** Note: additional arguments may be needed depending on models selected within the functions section.

| Argument | Name | Description |
|---|---|---|
| **Data objects** | | |
| `simulation_settings` | Simulation settings | A list of parameters governing the simulation time step settings |
| `demography` | Demography information | A tibble of demographic information for each individual (e.g., sex, socioeconomic status …) |
| `observation_times` | Serological survey settings | A tibble of observation times and biomarker for each individual |
| `foe_pars` | Force of exposure parameters | A 3D array providing the force of infection or vaccination for each exposure event, group and time |
| `biomarker_map` | Exposure to biomarker map | A table specifying the relationship between exposure types (*x*) and biomarker (*b*) |
| `model_pars` | Model parameters | A tibble of parameters needed for the antibody kinetics model, immunity model, observation model and the **draw_parameters** function |
| **Functions** | | |
| `exposure_model` | Exposure model | A function which will determine the probability that an individual is exposed to an exposure event which can be any event which leads to biomarker production. |
| `immunity_model` | Immunity model | A function which determines whether an exposure event is successful conditional on what the actual exposure is (vaccination, infection or re-infection) and the relevant factors. |
| `antibody_model` | Antibody model | A function that tracks antibody kinetics, or more broadly biomarker kinetics for each biomarker produced from successful exposure events. |
| `observation_model` | Observation model | A function which indicates how observed biomarker quantity Y are generated as a probabilistic function of the true, latent biomarker quantity *A.* |
| `draw_parameters` | Draw model parameters | A function to simulate parameters needed for the antibody model from `model_pars` |

**S2 Table. Description of `runserosim` outputs.**

| Output | Name | Description |
|---|---|---|
| exposure_histories | Exposure histories | An array of exposure histories across all individuals, time steps and exposure events |
| exposure_force | probability of an exposure event | An array of exposure probabilities across all individuals, time steps and exposure events. The exposure probability is the output of **exposure_model**. |
| exposure_probabilities | Probability of a successful exposure event | An array of the probability of a successful exposure event across all individuals, time steps and exposure events. The probability of a successful exposure event is the probability of exposure (output of **exposure_model**) multiplied by the probability of success (output of **immunity_model**) |
| biomarker_states | Biomarker states | True biomarker quantities for all individuals across all time steps and biomarker(s) |
| observed_biomarker_ states | Observed biomarker states | True and observed biomarker quantities at the observation time for the observed individuals and biomarker(s) |
| kinetics_parameters | Kinetics parameters | The simulated antibody kinetics parameters for all individual's successful exposure events. |

**S3 Table. Description of functions to plot `runserosim` outputs.**

| Name | Description |
|---|---|
| plot_subset_individuals_history | Plots biomarker quantities and exposure histories for a subset of individuals |
| plot_exposure_histories | Plots exposure histories across time for all individuals and exposure events; requires `exposure_histories_long` |
| plot_exposure_force | Plots the probability of an exposure event across time for all individuals and exposure events; requires `exposure_force_long` |
| plot_exposure_probabilities | Plots the probability of a successful exposure event across time for all individuals and exposure events; requires `exposure_probabilities_long` |
| plot_biomarker_quantity | Plots true biomarker quantities across all time steps for all individuals and biomarkers; requires `antibody_states` |
| plot_obs_biomarkers_one_sample | Plots observed biomarker quantities for all individuals and biomarkers; requires `observed_antibody_states` |
| plot_obs_biomarkers_paired_sample | Plots observed paired biomarker quantities for all individuals and biomarkers; requires `observed_antibody_states` |
| plot_exposure_model | Plots the probability of exposure over time for the provided exposure model |
| plot_antibody_model | Plots example trajectories of the provided antibody kinetics model |

**S4 Table. Names and descriptions of the ready-to-use exposure models included in serosim.**

| Name | Description |
|---|---|
| **Exposure Model** | |
| exposure_model_fixed | Probability of exposure for each time point is drawn directly from the `foe_pars` array. |
| exposure_model_simple_FOE | Probability of exposure ($1-e^{-\lambda}$) depends on the force of exposure ($\lambda$) at that time for that group. |
| exposure_model_dem_mod | Probability of exposure ($1-e^{-\lambda}$) depends on the force of exposure ($\lambda$) at the current time *t* for group *g* modulated by relevant demographic elements specified within the `dem_mod`. Within `dem_mod`, users can specify which demographic elements (including age) and how they affect the probability of exposure and by how much. |
| exposure_model_sir | Finds the probability of exposure governed by an SIR model with specified parameters for each exposure type and group combination. |
| exposure_model_gaussian_process | Generates an incidence curve (probability of infection per unit time) and associated parameters from a Gaussian Process model assuming that the covariance function (kernel) on time follows the squared exponential covariance function. |

**S5 Table. Names and descriptions of the ready-to-use immunity models included in serosim.**

| **Name** | **Description** |
|---|---|
| **Immunity Model** | |
| immunity_model_all_successful | Probability of success is 1 so all exposure events are successful immunologically. |
| immunity_model_vacc_only | This immunity model should only be used if all exposures are vaccination events. The probability of successful exposure(vaccination event) depends on the number of vaccines an individual has received prior to time *t* and whether an individual is eligible for vaccination. If the individual is under the maximum vaccinations allotted and eligible for vaccination then the probability of a successful exposure event is 1. |
| immunity_model_vacc_ifxn_simple | This immunity model should only be used with vaccines and natural infection. The probability of successful exposure for vaccination events depends on the number of vaccines an individual has received prior to time *t* and their current age. If the individual is under the maximum vaccinations allotted and is of an age eligible for vaccination then the probability of successful exposure is 1. The probability of a successful natural infection is dependent on the total number of infections that individual has experienced thus far. If the individual is under the maximum number of infections allotted then the probability of successful exposure event is 1. |
| immunity_model_ifxn_biomarker_prot | This immunity model should only be used for natural infection events. The probability of a successful exposure event is dependent on the individual's biomarker quantity at the time of exposure. User specified `biomakrer_prot_midpoint` and `biomarker_prot_width` within `model_pars` is used to calculate biomarker-mediated protection. |
| immunity_model_vacc_ifxn_biomarker_prot | This immunity model should be used if exposures represent vaccination and natural infection events. The probability of a successful vaccination exposure event depends on the number of vaccines received prior to time *t* while the probability of successful infection is dependent on the biomarker quantity at the time of exposure and the total number of successful infections prior to that point This immunity model combines **immunity_model_vacc_only** and **immunity_model_ifxn_biomarker_prot**. |

**S6 Table. Names and descriptions of the ready-to-use antibody models and model_pars files included in serosim.**

| Name | Description |
|---|---|
| **Antibody Model** | |
| antibody_model_monophasic | This monophasic antibody boosting-waning model assumes that for each exposure there is a boost and boost waning parameter. |
| antibody_model_biphasic | Biphasic antibody boosting-waning model with titer-dependent boosting. This model assumes that for each exposure there is a set of long-term boost, long-term boost waning, short-term boost, and short-term boost waning parameters. This model incorporates titer-ceiling effects by taking into account an individual's preexisting titer at the time of a new exposure event. |
| antibody_model_typhoid | Power law antibody boosting and waning model |
| **Model parameters** | |
| model_pars_README.csv | This file was used for the README file and the example within the paper. This model parameter tibble provides a template for simulations with 2 exposure events each containing the same biomarker, `antibody_model_monophasic`, `immunity_model_vacc_ifxn_simple`, `draw_parameters_random_fx` and `observation_model_continuous_noise`. |
| model_pars_cs1.csv | This file was used for case study 1 and provides a template for simulations with 2 exposure events each containing the same biomarker. Exposure 1 represents natural infection and exposure 2 represents vaccination. Users wishing to simulate a similar system simply need to input their desired parameter values into the mean, sd and distribution columns. This file includes all parameters needed for `antibody_model_biphasic`, `draw_parameters_random_fx_biomarker_dep`, `immunity_model_vacc_ifxn_biomarker_prot` and `observation_model_continuous_bounded_noise`. |
| model_pars_cs2.csv | This model_pars file was used for case study 2 and provides a template for simulations with 3 exposure events. Exposure 1 represents vaccination with 2 biomarkers, while exposures 2 and 3 represent natural infection boosting each biomarker individually. Users wishing to simulate a similar system simply need to input their desired parameter values into the mean, sd and distribution columns. This file includes all parameters needed for `antibody_model_biphasic`, `draw_parameters_random_fx_biomarker_dep`, `immunity_model_vacc_ifxn_biomarker_prot` and `observation_model_continuous_bounded_noise`. |

**S7 Table. Names and descriptions of the ready-to-use draw_parameters functions included in serosim.**

| Name | Description |
|---|---|
| draw_parameters_ fixed_fx | This function draws parameters directly from the distributions specified by `model_pars` for the antibody model with fixed effects. This function ensures that all individuals have the same parameters. |
| draw_parameters_ random_fx | This function draws parameters directly from `model_pars` for the antibody model with random effects. Parameters are drawn randomly from a distribution with mean and standard deviation specified within `model_pars`. |
| draw_parameters_ fixed_fx_biomarker_de p | This function adds biomarker quantity ceiling effects to the **draw_parameters_fixed_fx** function. Here, an individual's realized biomarker boost is dependent on their biomarker quantity at the time of the exposure event. |
| draw_parameters_ random_fx_biomarker_ dep | This function adds biomarker quantity ceiling effects to the previous **draw_parameters_random_fx** function. Here an individual's realized biomarker boost is dependent on their biomarker quantity at the time of the exposure event. |

**S8 Table. Names and descriptions of the ready-to-use observation models included in serosim.**

| Name | Description |
|---|---|
| **Observation Model** | |
| observation_model_continuous | This observation model observes the latent biomarker quantities given a continuous assay with no added noise. Therefore the observed biomarker quantity is simply given by the true latent biomarker quantity. |
| observation_model_continuous_bounded | This observation model observes the latent biomarker quantities given a continuous assay with user-specified lower and upper limits and no added noise. |
| observation_model_discrete | This observation model observes the latent biomarker quantities given a discrete assay with user-specified ranges within the `discrete` argument and no added noise. |
| observation_model_continuous_noise | This observation model observes the latent biomarker quantities given a continuous assay with added noise. The added noise represents assay variability and is done by sampling from a distribution with the latent biomarker quantity as the mean and the measurement error as the standard deviation. The observation standard deviation and distribution are defined within `model_pars` as the “obs_sd″ parameter. The user can also use the optional `sensitivity` and `specificity` arguments to account for assay sensitivity and specificity. False negatives are simulated by setting an observed quantity to 0 with probability `sensitivity`. False positives are simulated by drawing a random quantity from the bounded range for a true 0 biomarker quantity with probability 1-`specificity`. |
| observation_model_continuous_bounded_noise | This observation model observes the latent biomarker quantities given a continuous assay with user-specified lower and upper limits and added noise. The added noise represents assay variability and is done by sampling from a distribution with the latent biomarker quantity as the mean and the measurement error as the standard deviation. The observation standard deviation and distribution are defined within `model_pars` as the “obs_sd″ parameter. The user can also use the optional `sensitivity` and `specificity` arguments to account for assay sensitivity and specificity. False negatives are simulated by setting an observed quantity to the assay's lower bound with probability `sensitivity`. False positives are simulated by drawing a random quantity from the bounded range for a true 0 biomarker quantity with probability 1-`specificity`. |
| observation_model_discrete_noise | This observation model observes the latent biomarker quantities given a discrete assay with user-specified ranges within discrete and added noise. The added noise represents assay variability and is done by sampling from a distribution with the latent biomarker quantity as the mean and the measurement error as the standard deviation. The observation standard deviation and distribution are |

| | |
|---|---|
| | defined within model_pars as the “obs_sd” parameter. The user can also use the optional sensitivity and specificity arguments to account for assay sensitivity and specificity. False negatives are simulated by setting an observed quantity to the assay's lower bound with probability sensitivity. False positives are simulated by drawing a random quantity from the bounded range for a true 0 biomarker quantity with probability 1-specificity. |
